# Antibody Responses 3-5 Months Post-Vaccination with mRNA-1273 or BNT163b2 in Nursing Home Residents

**DOI:** 10.1101/2021.08.17.21262152

**Authors:** Jessica A. Breznik, Ali Zhang, Angela Huynh, Matthew S. Miller, Ishac Nazy, Dawn M. E. Bowdish, Andrew P. Costa, for the COVID-in-LTC Study Group

## Abstract

Nursing home residents often fail to mount robust responses to vaccinations and recent reports of breakthrough infections, particularly from variants of concern, raise questions about whether vaccination regimens elicit a sufficient humoral immune response or if booster doses are warranted. We examined SARS-CoV-2 antibody levels and neutralizing capacity in nursing home residents 3-5 months after 2 doses of mRNA-1273 or BNT163b2 vaccination as per recommended schedules.

Nursing home residents were recruited from eight long-term care homes in Ontario, Canada, between March and July 2021. Antibody levels and neutralization capacity from a previously published convalescent cohort were used as a comparator. Serum SARS-CoV-2 IgA/G/M against spike (S) protein and its receptor-binding domain (RBD) were measured by validated ELISA, with assay cut-off at the mean and 3 standard deviations of a pre-COVID-19 population from the same geographic region. Antibody neutralization was measured against the wild-type strain of SARS-CoV-2 and the beta variant of concern (B.1.351).

No neutralizing antibodies were detected in ∼20% of residents to the wild-type virus (30/155; 19%) or beta variant (27/134; 20%). Residents that received BNT163b2 had a ∼4-fold reduction in neutralization to the wild-type strain, and a ∼2-fold reduction in neutralization to the beta variant relative to those who received mRNA-1273.

Current mRNA SARS-CoV-2 vaccine regimens may not have equivalent efficacy in nursing home residents. Our findings imply that differences in the humoral immune response may contribute to breakthrough infections, and suggest that consideration of the type of vaccine administered to older adults will have a positive impact on the generation of protective immunity.

## Introduction

Nursing home residents in Ontario, Canada, were prioritized for vaccination with mRNA vaccines from Moderna (mRNA-1273) or Pfizer (BNT163b2) in December 2020-January 2021, which significantly reduced the high morbidity and mortality due to COVID-19^1^. Yet nursing homes residents often fail to mount robust responses to vaccinations^2^ and recent reports of breakthrough infections, particularly from variants of concern, raise questions about whether vaccination regimens elicit a sufficient humoral immune response or if booster doses are warranted.

## Methods

We examined SARS-CoV-2 antibody levels and neutralizing capacity in nursing home residents 3-5 months after 2 doses of mRNA-1273 or BNT163b2 vaccination as per recommended schedules. Nursing home residents were recruited from eight long-term care homes in Ontario, Canada, between March and July 2021. Antibody levels and neutralization capacity from a previously published convalescent cohort were used as a comparator^2^. All protocols were approved by the Hamilton Integrated Research Ethics Board, and informed consent was obtained.

Venous blood was drawn in anti-coagulant-free vacutainers for isolation of serum. Serum SARS-CoV-2 IgA/G/M against spike (S) protein and its receptor-binding domain (RBD) were measured by validated ELISA, with assay cut-off at the mean and 3 standard deviations of a pre-COVID-19 population from the same geographic region^3^. Data are reported as a ratio of observed optical density (OD) to the determined assay cut-off OD, with ratios above 1 considered positive. Neutralization capacity of these antibodies was assessed by cell culture assays with live SARS-CoV-2 virus, with data reported as geometric microneutralization titers at 50% (MNT_50_) which ranged from below detection (MNT_50_ = 10) to MNT_50_ = 1280^3^. Antibody neutralization was measured against the wild-type strain of SARS-CoV-2 and the beta variant of concern (B.1.351). The beta variant was obtained through BEI Resources, NIAID, NIH: SARS-Related Coronavirus 2, Isolate hCoV-19/South Africa/KRISP-K005325/2020, NR-54009, contributed by Alex Sigal and Tulio de Oliveira.

Differences between antibody levels and neutralization in individuals that received mRNA-1273 or BNT163b2 were assessed by chi-square of independence (proportions), Kruskal-Wallis test (median), and student’s t-test (mean). All statistical analyses were conducted using SAS 9.4 (SAS Institute Inc.).

## Results

The majority of residents (97.1%) produced antibodies to the S protein post vaccination; however, fewer residents (87.68%) produced IgG to the RBD domain (**Table**). Residents who received mRNA-1273 had higher levels of IgG S protein (mRNA-1273: 2.9, IQR 2.5-3.1;) and IgG RBD (mRNA-1273: 2.5, IQR 1.7-3.0;) than those who received BNT163b2 (Ig Spike: BNT163b2: 2.5, IQR 1.5-3.1; p=0.015, Ig RBD: BNT163b2: 1.5, IQR 0.7-2.6; p<0.0001). Participants who had been vaccinated with BNT163b2 had median values of both Ig Spike and RBD that were lower than the median values of a cohort of convalescent individuals. There were no differences between vaccine groups with respect to IgM/A to either S protein or RBD. No neutralizing antibodies were detected in ∼20% of residents to the wild-type virus (30/155; 19%) or beta variant (27/134; 20%). Residents that received BNT163b2 had a ∼4-fold reduction in neutralization to the wild-type strain, and a ∼2-fold reduction in neutralization to the beta variant relative to those who received mRNA-1273 (**Figure**).

**Table.**
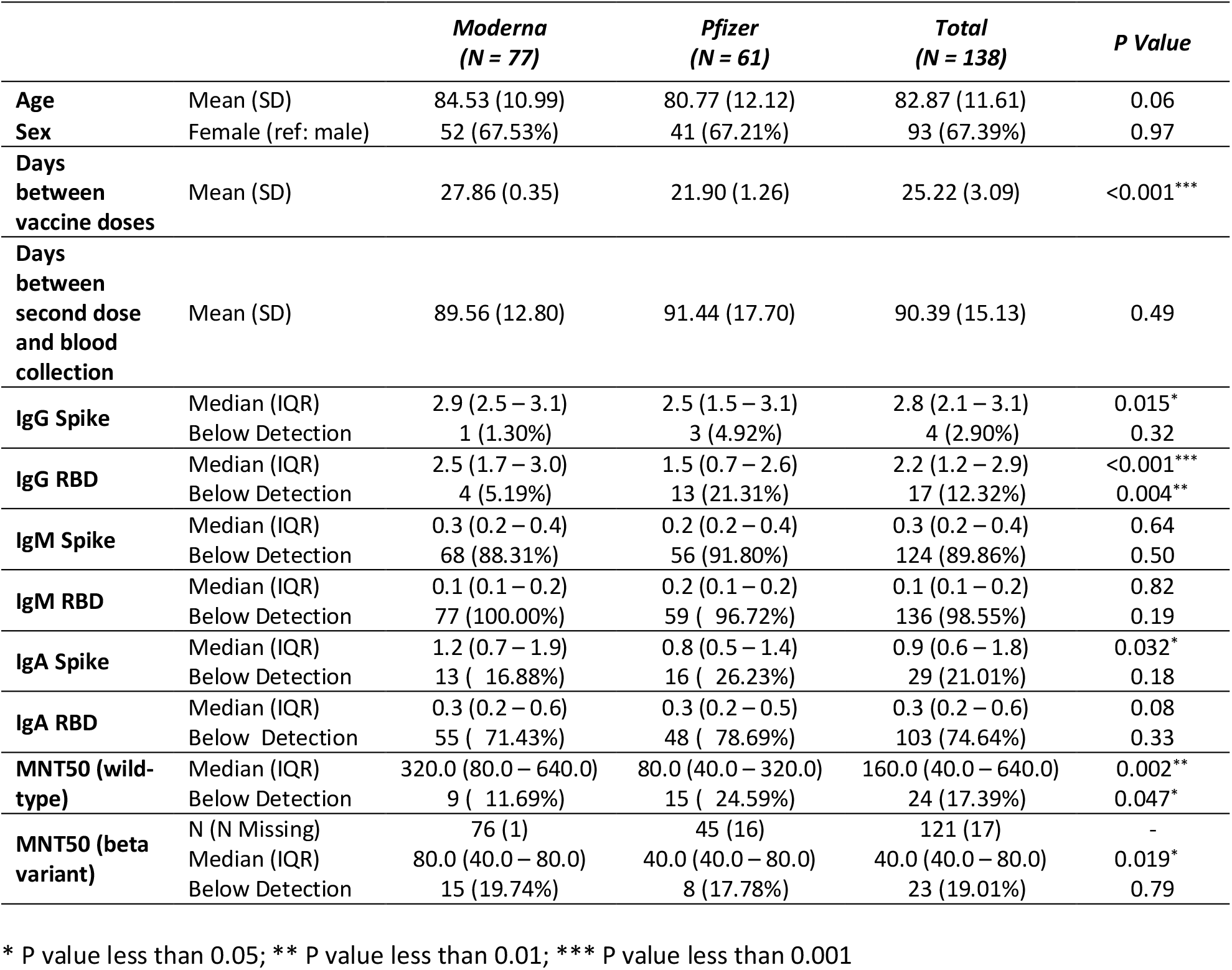
Antibody levels and virus neutralization capacity 60-130 days post-vaccination in nursing home residents.

**Figure.**
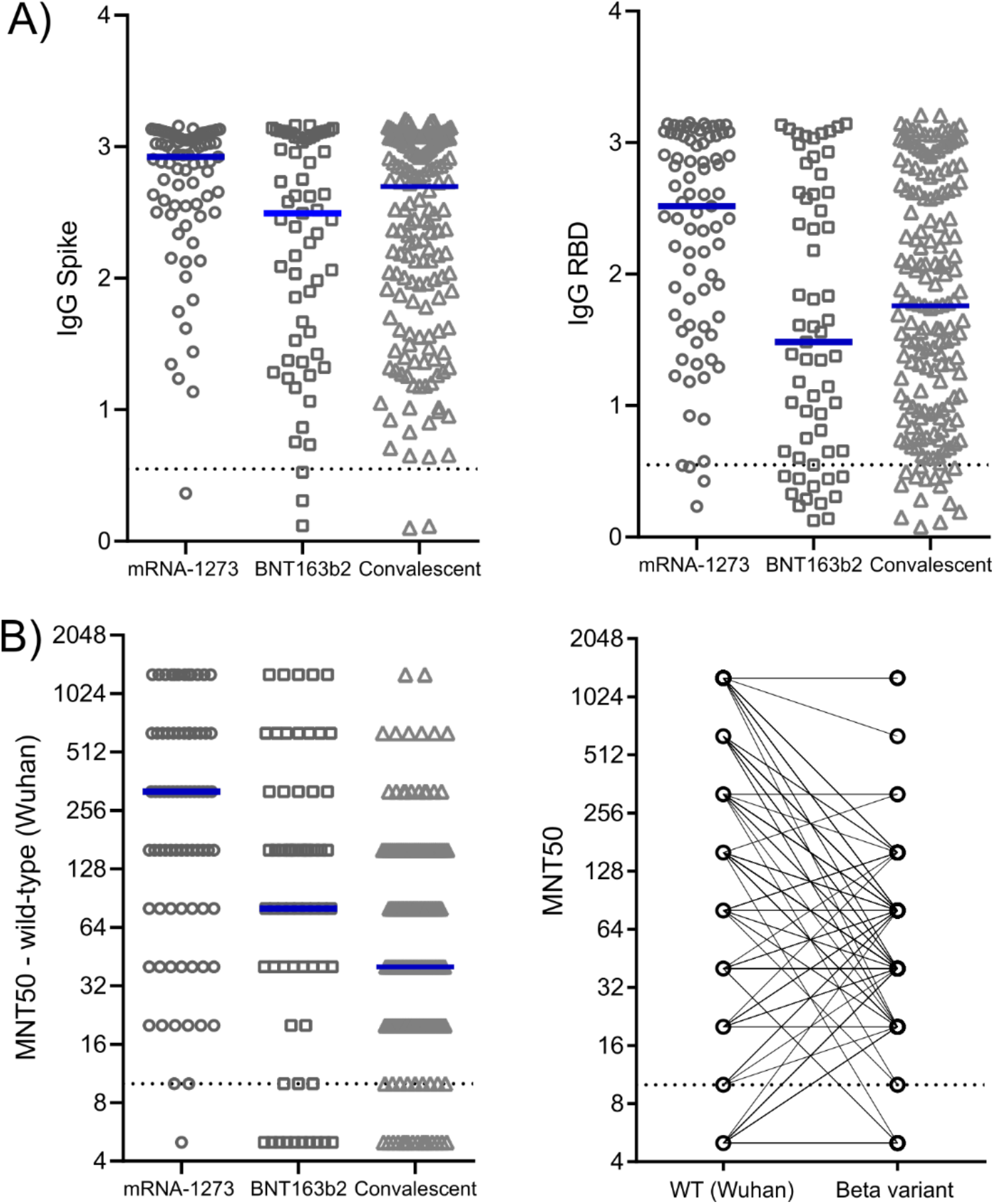
IgG antibody titers (A) and virus neutralization titers (B) by mRNA vaccine type 3-5 months post-vaccination in nursing home residents. Blue bars indicate median values. Dotted lines represent the level of detection. Microneutralization titres (MNT) are visualized as a log scale. Individuals who did not have any detectable neutralization capacity (Wuhun: n = 9, mRNA-1273 and n=15 BNT163b) were assigned a value of 5.

## Discussion

Two does of vaccine failed to elicit any antibody-mediated protective immunity in ∼20% of nursing home residents. These data align with recent observations of decreased antibody production and/or neutralization after BNT162b2 vaccination in nursing home residents compared to healthy young individuals^4-6^. In addition, we found that vaccination against SARS-CoV-2 with mRNA-1273 elicited a stronger humoral response compared to BNT162b2, with greater circulating IgG and neutralization antibody titers 3 - 5 months after vaccination. The mRNA-1273 vaccine contains a higher dose of mRNA, which may imply that a higher dose is beneficial to generate protective immunity in nursing home residents.

## Data Availability

Data are not available without prior approval from the Hamilton Integrated Research Ethics Board (HiREB).

